# TBtypeR: Sensitive detection and sublineage classification of low-frequency *Mycobacterium tuberculosis* complex mixed infections

**DOI:** 10.1101/2024.06.12.24308870

**Authors:** Jacob E Munro, Anna K Coussens, Melanie Bahlo

## Abstract

Mixed infections comprising multiple *Mycobacterium tuberculosis* Complex (MTBC) strains are observed in populations with high incidence rates of tuberculosis (TB), yet the difficulty to detect these via conventional diagnostic approaches has resulted in their contribution to TB epidemiology and treatment outcomes being vastly underrecognised. In endemic regions, detection of all component strains is crucial for accurate reconstruction of TB transmission dynamics. Currently available tools for detecting mixed infections from whole genome sequencing (WGS) data have insufficient sensitivity to detect low-frequency mixtures with less than 10% minor strain fraction, leading to a systematic underestimation of the frequency of mixed infection. Our R package, TBtypeR, identifies mixed infections from whole genome sequencing by comparing sample data to an expansive phylogenetic SNP panel of over 10,000 sites and 164 MTBC strains. A statistical likelihood is derived for putative strain mixtures based on the observed reference and alternative allele counts at each site under the binomial distribution. This provides robust and high-resolution sublineage classification for both single- and mixed-infections with as low as 1% minor strain frequency. Benchmarking with simulated *in silico* and *in vitro* mixture data demonstrates the superior performance of TBtypeR over existing tools, particularly in detecting low frequency mixtures. We apply TBtypeR to 5,000 MTBC WGS from a published dataset and find a 6-fold higher rate of mixed infection than existing methods. The TBtypeR R package and accompanying end-to-end Nextflow pipeline are available at github.com/bahlolab/TBtypeR.

## Introduction

Tuberculosis (TB) stubbornly remains the leading cause of infectious morbidity and mortality, with over 1 million annual deaths and 10 million annual cases for the last two decades^1^. Global eradication of TB is hampered by the proliferation of drug-resistant *Mycobacterium tuberculosis* (*Mtb*) strains, the lack of an effective vaccine for adults and our inability to stop transmission. TB patients infected with a mixture of one or more *Mtb* strains (mixed infection) have been reported in endemic regions across multiple studies^2–6^, with wide-ranging prevalence estimates from 0.4% to 57.1%^6^. Such mixed infections are associated with poorer treatment outcomes^7^. Whilst the occurrence of TB mixed infections has been established for some time, the contribution and importance of this phenomenon to local and global epidemiology of TB remains substantially underexplored. Characterising the extent of mixed infections, and thus re-infection, within populations is crucial for our understanding of the natural history of TB pathogenesis. Furthermore, inadequate detection of mixed infection impedes efforts to confirm transmission events, model transmission dynamics and confounds the classification of recurrent TB as relapses or reinfections, collectively hindering the effective design and implementation of TB control programs in endemic regions^8^.

The contribution of TB mixed infection has been largely ignored because the highly clonal genetic structure of *Mtb*, its low mutation frequency and absence of on-going horizontal gene transfer, resulting in significantly less genetic diversity amongst isolates than other bacterial pathogens^9^. Consequently, the most commonly used routine diagnostic lab tests in high-burden settings for genetically typing *Mtb* strains (with restriction fragment length polymorphism [RFLP] or spacer oligonucleotide [spoligo] typing) use repeat elements that lack sensitivity to detect the majority of mixed infections, except when from highly divergent strains. Whilst whole genome sequence-based typing now enables strain sublineage classification, existing analytical pipelines lack sufficient sensitivity to genotype low frequency isolate contributions.

The *Mycobacterium tuberculosis* complex (MTBC) comprises five genetically close species of *Mycobacterium* known to cause TB and has been divided into 12 monophyletic lineages based on genomic phylogenetic reconstruction: Lineage 1 (Indo-Oceanic *Mtb*), Lineage 2 (East-Asian *Mtb*), Lineage 3 (East-African-Indian *Mtb*), Lineage 4 (Euro-American *Mtb*), Lineage 5 (West-African 1 *M. africanum*), Lineage 6 (West-African 2 *M. africanum*), Lineage 7 (Ethiopian *Mtb*), Lineage 8 (African Great Lakes *Mtb*), Lineage 9 (East African *M. africanum*), La1 (*M. bovis*), La2 (*M. caprae*), and La3 (*M. orygis*)^10–13^. Several efforts have been made to identify single nucleotide polymorphism (SNP) markers (known as SNP “barcodes”) unique to specific MTBC lineages and sublineages (monophyletic) for use in strain identification and classification^10–12,14,15^, replacing the less-precise RFLP and spoligotyping.

Multiple informatic tools have been developed to detect mixed samples from MTBC WGS data over the last decade, all of which rely on analysing the reference and alternate (B) allele counts at SNPs that differ between strains in the mixture WGS, with the B-allele frequency at each site approximating the mixture frequency. The tools fall into two general classes: (1) barcode-based tools, which rely on a set of known phylogenetically defined SNP markers, and (2) barcode-free tools, which rely only on the observed WGS data to make mixture predictions. Barcode-based tools have the advantage that they are able to identify the specific sublineage of mixture components, whilst barcode-free tools have the advantage that they can identify mixtures between novel or closely related strains that are not present in the barcode.

Two key barcode-free tools are MixInfect^16^ and SplitStrains^17^. MixInfect was the first tool developed and employs a method based on calling variants with BCFtools^18^ followed by Bayesian Gaussian mixture modelling of the B-allele frequency to estimate the minor strain fraction. MixInfect can reliably identify component mixed infections with ≥10% minor strain fraction (sensitivity ≥ 0.92) but fails at 5% minor strain fraction (sensitivity = 0.33). SplitStrains similarly uses a Gaussian mixture model and classifies mixed and non-mixed samples with a likelihood ratio test. SplitStrains is also reliable at ≥10% minor strain fraction (sensitivity ≥ 0.92) but fails at 5% minor strain fraction (sensitivity = 0.08) with default parameters.

Barcode-based tools include TBProfiler^19^, Fastlin^20^ and QuantTB^21^. TBProfiler uses a phylogenetic barcode consisting of 1,101 SNPs covering 125 MTBC phylotypes (as of v5.0.1) and is capable of detecting mixed infections. This functionality was not described in the original manuscript, and the process used to detect mixed infection as well as the lower limits of detection are not documented. Fastlin uses the same phylogenetic SNP barcode as TBProfiler, however the SNPs are converted to k-mers of 25 base pairs by adding flanking sequence from the reference genome which are then used to rapidly scan sequencing read files without requiring alignment to a reference genome. Fastlin estimates the relative abundance of mixed strains based on the median count of all detected k-mers associated with each specific strain. The authors do not provide advice on the minimum minor strain fraction Fastlin is capable of detecting. Finally, QuantTB uses a non-phylogenetic SNP barcode constructed from 5,637 MTBC genome assemblies aligned to the H37Rv reference genome, yielding 2,167 strains and 244,181 SNP sites. Sample variants are first called at the barcode sites, after which an iterative algorithm is applied to select the strain which explains the most SNPs in the sample of interest until a threshold is met. The minor strain fraction is then estimated by averaging the B-allele frequency of SNPs unique to each strain identified. The authors demonstrated that QuantTB is able to identify mixtures at 10% minor strain fraction but did not assess performance below 10% minor strain fraction.

Here we describe our tool, TBtypeR, an easy-to-use R package and accompanying Nextflow pipeline, which has been designed for sensitive and accurate detection of low-frequency mixed infections with reliable detection of mixtures down to 2.5% minor strain fraction. We have performed thorough benchmarking of the existing tools for detection of mixed infection in MTBC WGS data, show the minimum minor strain fraction each tool is capable of detecting, illustrate the overall strengths and weaknesses of each approach and demonstrate that TBtypeR identifies a 6-fold higher rate of mixed infection in a published WGS dataset than conventional methods.

## Materials and Methods

### TBtypeR development

TBtypeR is an R^22^ package for mixed strain identification from WGS data based on a phylogenetic SNP barcode. The included SNP barcode consists of 10,903 sites covering 164 MTBC phylotypes (lineages and sublineages) compiled from Napier et al^10^, Zwyer et al^11^, Thawornwattana et al^14^, Coscolla et al^12^ and Shuaib et al^15^ (Table 1). The barcode is designed to be easily extended or replaced by users, allowing the tool to be run against emergent strains or with non-MTBC species. Input data for TBtypeR is provided as a multi-sample Genomic Data Structure (GDS) format^23^ or Variant Call Format^24^ (VCF) file. TBtypeR identifies strain mixtures by maximising the joint binomial likelihood across observed SNP sites using an iterative greedy search algorithm. At each iteration, TBtypeR identifies the best candidate strain to add to the mixture model and then optimises both the mixture proportions and the background error rate by maximising the joint likelihood. Statistical significance is assessed by both a permutation test and a Wilcoxon rank sum test with false discovery rates controlled by the Benjamini-Hochberg procedure. TBtypeR is provided as both a stand-alone R package and as part of a comprehensive Nextflow^25^ pipeline, TBtypeNF, which additionally performs FASTQ preprocessing with fastp^26^, read alignment with BWA-MEM^27^, variant calling with BCFtools^18^, and quality control (QC) report generation using SAMtools^18^, mosdepth^28^ and multiQC^29^. An alternative “Fast” workflow, FastTBtypeNF, was implemented in Nextflow by using Fastlin with a modified barcode to rapidly generate allele counts for use by TBtypeR, and thus avoiding the time-consuming alignment, variant calling, and QC steps. These three variants provide options for user customisation (TBtypeR), ease-of-use and convenience (TBtypeNF), and speed and cost-effectiveness (FastTBtypeNF).

**Table 1:**
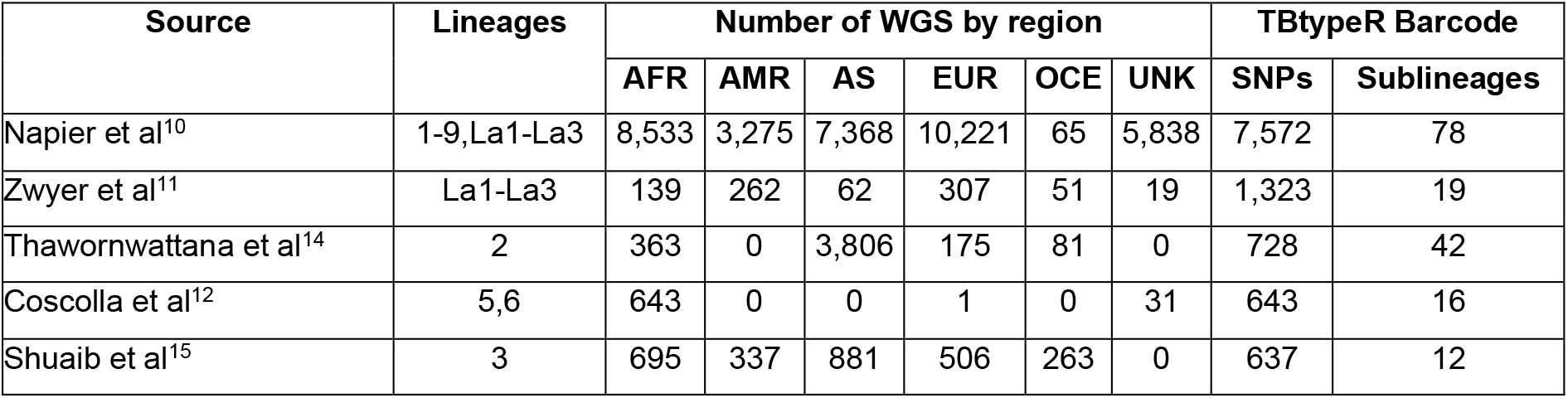
Data sources for the TBtypeR SNP Barcode. Region Abbreviations; AFR: Africa, AMR: Americas, AS: Asia, EUR: Europe, OCE: Oceania, UNK: unknown region. Sources are ordered by publication date. SNPs identified in multiple sources are counted once and attributed to the first published source.

| Source | Lineages | Number of WGS by region |  |  |  |  |  | TBtypeR Barcode |  |
| --- | --- | --- | --- | --- | --- | --- | --- | --- | --- |
|  |  | AFR | AMR | AS | EUR | OCE | UNK | SNPs | Sublineages |
| Napier et al<sup>10</sup> | 1-9,La1-La3 | 8,533 | 3,275 | 7,368 | 10,221 | 65 | 5,838 | 7,572 | 78 |
| Zwyer et al<sup>11</sup> | La1-La3 | 139 | 262 | 62 | 307 | 51 | 19 | 1,323 | 19 |
| Thawornwattana et al<sup>14</sup> | 2 | 363 | 0 | 3,806 | 175 | 81 | 0 | 728 | 42 |
| Coscolla et al<sup>12</sup> | 5,6 | 643 | 0 | 0 | 1 | 0 | 31 | 643 | 16 |
| Shuaib et al<sup>15</sup> | 3 | 695 | 337 | 881 | 506 | 263 | 0 | 637 | 12 |

### Benchmarking Datasets

Three datasets were used to benchmark the tools. The first is the “*in vitro* duos” mixture dataset published along with the MixInfect tool^16^, which consists of 48 mixtures derived from mixing DNA extracted from pure *Mtb* cultures, with 12 each of minor strain fraction equal to 0% (unmixed), 5%, 10% and 30%. The second and third datasets are the “*in silico* duos” and “*in silico* trios” datasets which were constructed by mixing sequencing reads from a subset of 701 MTBC WGS data published by the CRyPTIC Consortium^30^. Samples were selected based on the following criteria: sequencing platform equal to “Illumina HiSeq 2500”, median depth ≥ 100, fastp percent “pass” reads ≥ 95%, SAMtools percent mapped reads ≥ 95%, read length ≥ 290 bp and mean SNP B-allele frequency < 0.0007 (determined empirically as indicative of a non-mixed sample). The “*in silico* duos” dataset consisted of 50 samples at each combination of coverage (20x, 40x, 60x, 80x and 100x) and minor strain fraction (1%, 2.5%, 5%, 10%, 25% and 50%), giving 1500 mixed samples and a further 1500 matched unmixed samples using the same major strain for a total of 3000 simulated samples. The “*in silico* trios” dataset consisted of 250 unmixed samples, 250 matched mixtures of 2 strains with a 1:1 ratio, and 250 matched mixtures of 3 strains with a 3:2:1 ratio with equal numbers of samples with coverage 20x, 40x, 60x, 80x and 100x. Mixtures were generated by downsampling FASTQ files to the desired coverage with SeqKit^31^ and then concatenating the reads into a single FASTQ file. Samples in each simulated mixture were chosen semi-randomly from the pool of 701, avoiding combinations where there were fewer than 10 SNPs different between component samples. Source code for generating the “*in vitro*” and “*in silico*” mixture datasets is provided in the TBtypeR repository.

### Benchmarking

In order to compare the performance of TBtypeR to other published tools, Nextflow workflows were implemented for each of Fastlin, TBProfiler, SplitStrains, QuantTB and MixInfect and are included in TBtypeR repository. Docker containers were created based on Fastlin (v0.2.3, installed via Bioconda), TBProfiler (v5.0.1, installed via Bioconda), SplitStrains (GitHub commit “f936cb2”, dependencies installed via Conda), QuantTB (v1.01, installed via Conda) and MixInfect (GitHub commit “79a7315”, dependencies installed via Conda). Fastlin, TBProfiler and QuantTB take FASTQ files directly as input so were straightforward to implement as Nextflow processes. SplitStrains required preprocessing of FASTQ files which was implemented based on the “gcs.sh” and “runSplitStrains.sh” scripts available on GitHub (https://github.com/WGS-TB/SplitStrains). MixInfect also required preprocessing which was implemented according to the description provided in Sobkowiak et al^16^. Each tool was used with default arguments unless otherwise specified. Since TBProfiler, Fastlin and QuantTB make use of SNP barcodes, these tools were benchmarked with both the provided barcode as well as one derived from the TBtypeR barcode to isolate performances differences based on SNP barcode alone.

In order to assess the runtime of each tool, 100 samples from the “in silico duos” dataset with simulated coverage of 60x were selected, with equal numbers of unmixed and 50% minor strain fraction samples. Benchmarks were performed on a SLURM based HPC and run on nodes with Intel® Xeon® Gold 6342 CPUs and repeated 5 times each. CPU time was calculated by multiplying the percentage of CPU utilisation by the process real-time in seconds as reported by the Nextflow process trace. The median CPU time was taken across the 5 replicates for each sample, and a mean value was calculated across all samples for each tool.

Data analyses and visualisations were performed with R^22^, Posit Workbench, Rmarkdown and the tidyverse suite of packages^32^. Source code for running benchmarks and generating figures and tables is provided in the TBtypeR repository.

## Results

Each tool was assessed with a suite of benchmarking tests on the “*in vitro*” and “*in silico*” mixtures datasets and the performance ranked by the Mathew’s correlation coefficient^33^ (MCC), the mean absolute error (MAE) of minor strain fraction prediction, and the runtime per sample (Table 2). The MCC was preferred over other measures of binary classification performance, such as the F_1_ score, as it is robust on unbalanced datasets and invariant to class-swapping^34^. TBtypeR performed best in six of the seven overall benchmark categories, while Fastlin performed best in the runtime benchmark. An overall performance score was calculated and used to rank the tools as follows: 1) TBtypeR; 2) Fastlin; 3) TBProfiler; 4) QuantTB; 5) MixInfect; and 6) SplitStrains. Replacement of the default SNP barcodes with the TBtypeR barcode improved the overall performance of the other barcode-based tools (Fastlin, TBProfiler and QuantTB). For QuantTB the accuracy of minor strain fraction estimation was substantially improved, however the overall binary classification performance was somewhat lower when using the TBtypeR barcode. Interestingly, the alternative FastTBtypeNF workflow was found to slightly outperform the default TBtypeNF workflow while being eleven times faster to run, suggesting that Fastlin’s k-mer counting approach may be more accurate than traditional alignment for strain assignment.

**Table 2:** Overall performance and ranking of each tool on the “in vitro” and “in silico” benchmark datasets. “MCC” is the Mathews correlation coefficient (higher values are better), “MAE” is the mean absolute error in assigned minor strain fraction percentage (lower values are better) and “CPU sec.” is the average number of CPU seconds required to process a sample with a coverage of 60x (lower values are better). The overall “Score” is the mean of the linear rescaling of each metric to the interval [0,1], where 0 is assigned to the worst performer and 1 to the best performer. Tools are ordered from best to worst overall score, with best performers in each column indicated with bold font and worst performers in each column indicated with an underline. “+BC” in tool name indicates replacement of the tool’s default SNP barcode with the TBtypeR barcode.

| Tool | Score | *In vitro* duos<br>(n=48) |  | *In silico* duos<br>(n=3000) |  | *In silico* trios<br>(n=500) |  | Runtime<br>(n=100) |
| --- | --- | --- | --- | --- | --- | --- | --- | --- |
|  |  | MCC | MAE (%) | MCC | MAE (%) | MCC | MAE (%) | CPU sec. |
| TBtypeR (FastTBtypeNF) | **0.985** | **0.944** | **1.65** | 0.840 | **0.88** | 0.796 | **1.80** | 020.5 |
| TBtypeR (TBtypeNF) | 0.951 | **0.944** | 1.67 | **0.864** | 1.14 | **0.827** | 1.95 | 222.6 |
| Fastlin (+BC) | 0.766 | 0.775 | 6.24 | 0.707 | 3.11 | 0.810 | 2.62 | 004.3 |
| TBProfiler (+BC) | 0.762 | 0.713 | 2.18 | 0.551 | 1.96 | 0.763 | 2.23 | 267.6 |
| Fastlin | 0.746 | 0.856 | 3.07 | 0.575 | 2.52 | 0.652 | 3.79 | **003.8** |
| TBProfiler | 0.652 | 0.602 | 3.66 | 0.539 | 2.47 | 0.642 | 3.49 | 249.2 |
| QuantTB (+BC) | 0.489 | 0.447 | 6.40 | <u>0.431</u> | 3.49 | 0.534 | 4.19 | 245.0 |
| QuantTB | 0.418 | 0.612 | <u>9.97</u> | 0.493 | <u>7.23</u> | 0.771 | 4.00 | 284.8 |
| MixInfect | 0.315 | 0.558 | 5.52 | 0.542 | 5.22 | <u>-0.283</u> | <u>6.41</u> | 414.7 |
| SplitStrains | <u>0.270</u> | <u>0.000</u> | 7.23 | 0.549 | 2.06 | 0.000 | 5.39 | <u>834.6</u> |

The performance of SplitStrains was notably inconsistent across these benchmarks. In the “*in vitro* duos” benchmark, specificity was low (0) and sensitivity was high (1), with all samples predicted as mixed. In contrast, in the “*in silico* duos” benchmark, specificity was high (0.99) and sensitivity was low (0.48). We note that the same *in vitro* dataset was used to benchmark SplitStrains in the original publication as has been used here, however with different results (specificity = 1, sensitivity = 0.86). Despite our best efforts to match the software versions, parameters, and data processing of the original publication, and reaching out to the authors for guidance, we were not able to reproduce these results with our Nextflow implementation.

The effects of sequencing coverage, minor strain fraction and pairwise SNP distance were explored for each of the tools on the “*in silico* duos” dataset using the MCC measure (Figure 1A). TBtypeR, Fastlin and SplitStrains, show improving performance with increased sequencing coverage as expected, while QuantTB and TBProfiler showed no trend, and MixInfect unexpectedly performed worse as sequencing coverage increased. Mixed infections are more difficult to detect when the component strains are genetically similar as there are few informative SNP sites on which to make predictions. Accordingly, the barcode-based tools (TBtypeR, Fastlin, TBProfiler and QuantTB) showed worst performance when the SNP distance between component strains was lowest. However, TBtypeR still outperformed the two barcode-free tools (MixInfect and SplitStrains) at the lowest SNP distance bin, whilst these two performed better than the other barcode-based tools at this SNP distance. The minor strain fraction had the largest effect on the performance of all tools, with performance dropping off at minor strain fractions below 10%. TBtypeR performed best at low minor strain fractions, with reliable performance (MCC > 0.7) maintained down to 2.5% minor strain fraction, followed by MixInfect down to 5% minor strain fraction, and the remaining tools down to 10% minor strain fraction. All tools, except QuantTB and MixInfect, maintained high specificity (>0.95) across the benchmark so decreased MCC was driven primarily by decreased sensitivity (Figure S1). At 1% minor strain fraction TBtypeR maintained an MCC of 0.63 while all other tools were effectively no better than a random guess classifier with MCC close to 0 (0.0-0.05). TBtypeR yielded the highest MCC across the majority of tested points (14/16) and was only slightly outperformed by SplitStrains when the minor strain fraction was greater than or equal to 25%.

**Figure 1:**
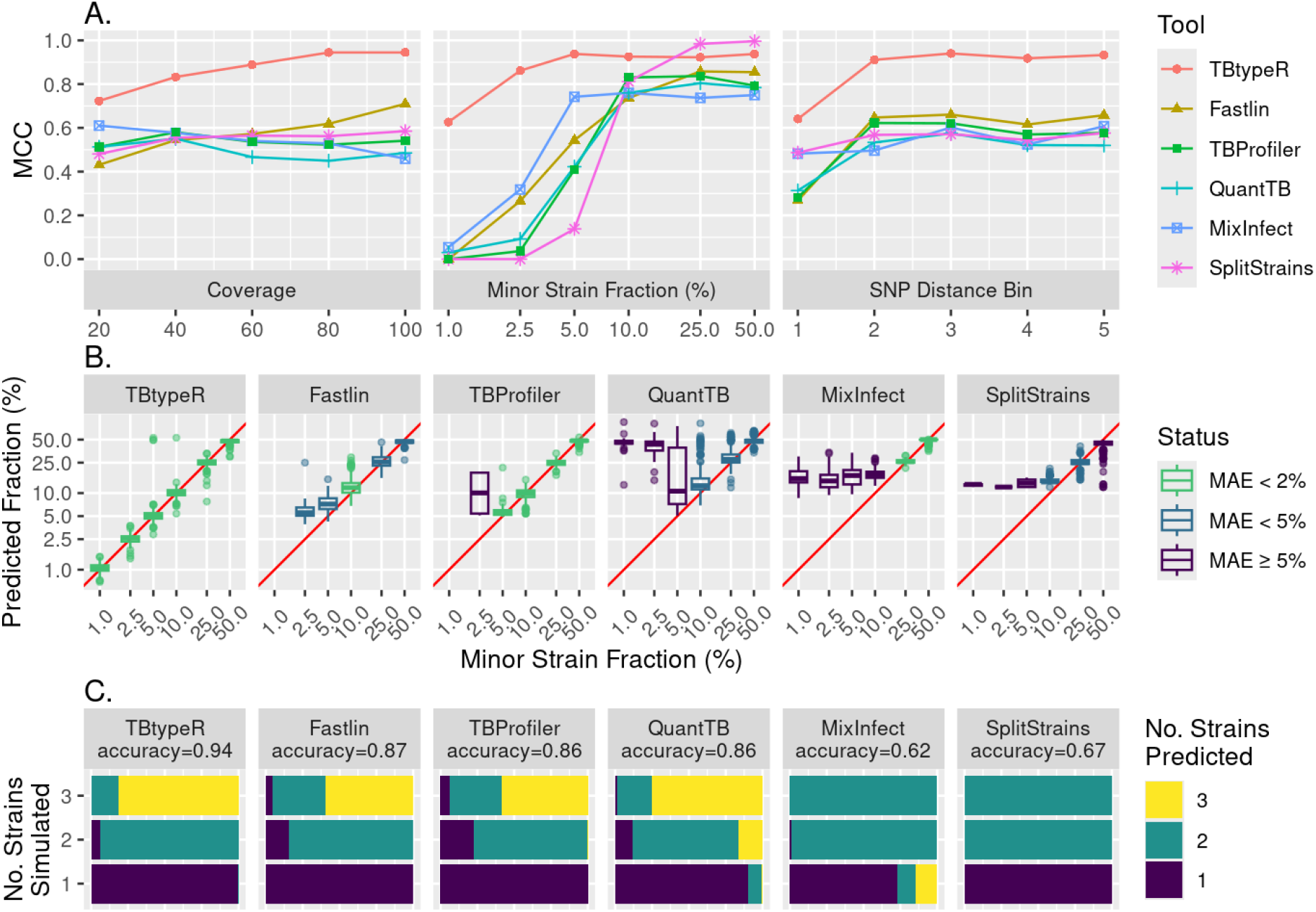
Performance measures of each tool on the “in silico” datasets. A) Mathews correlation coefficient (MCC) is plotted against simulated coverage, minor strain fraction, and SNP distance bin for the “in silico duos” dataset. Distance bins were derived by dividing the pairwise SNP distance between the major and minor strain into 5 bins of equal counts, with means and ranges as follows: bin 1: 67 (10-114); bin 2: 147 (115-180); bin 3: 187 (181-193); bin 4: 202 (194-216); and bin 5: 317 (217-432). The x-axis of minor strain fraction is log-2 scaled for improved resolution. B) Boxplots showing the distribution of predicted minor strain fraction for each simulated value across each tool in the “in silico duos” dataset. The red line corresponds to the function y=x. Axes are log-2 scaled for improved resolution and boxplots are coloured by MAE (the mean absolute error in assigned minor strain fraction percentage). Absent boxplots for TBProfiler and Fastlin are due to no mixture predictions being made at 1% minor strain fraction by these tools. C) Accuracy of each tool in predicting simulated mixtures of either one, two or three strains in the “in silico trios” dataset. The length and colour of the bars represents the proportion of samples predicted to have the specified number of strains.

In addition to best binary classification performance, TBtypeR was found to have the highest accuracy in minor strain fraction estimation, particularly at low minor strain fraction, maintaining a median absolute error (MAE) of less than 2% minor strain fraction, down to minor strain mixtures of 1% (Figure 1B). Next best was Fastlin, able to maintain a MAE less than 5% minor strain fraction down to minor strain mixtures of 2.5%, followed by TBProfiler maintaining a MAE less than 2% minor strain fraction down to minor strain mixtures of 5%. SplitStrains and QuantTB performed poorly, maintaining a MAE of less than 5% minor strain fraction down to minor strain mixtures of only 10%, and MixInfect performed worst maintaining a MAE of less than 2% minor strain fraction down to minor strain mixtures of only 25%.

The ability of each tool to identify mixtures of three strains (trios) was assessed with the “*in silico* trios” dataset and measured by the accuracy of assigning equal numbers of one-, two- and three-strain mixtures (Figure 1C). Here TBtypeR also performed best, correctly assigning the highest proportion of mixtures with an accuracy of 0.94, followed by Fastlin with an accuracy of 0.87, TBProfiler and QuantTB with accuracies of 0.86, SplitStrains with an accuracy of 0.67 and MixInfect with an accuracy of 0.62. It should be noted that as implemented SplitStrains is not able to predict three-strains mixtures as the mixture number needs to be provided a priori and is not estimated from the data. QuantTB and MixInfect were the only tools to make erroneous predictions of three-strain mixtures.

MTBC strain sublineage assignment was compared between TBtypeR, TBProfiler and Fastlin on 690 non-mixed samples downsampled to 100x coverage (Table S1). The QuantTB barcode is not phylogenetic and does not use standard nomenclature for MTBC sublineages so strain assignment could not be meaningfully compared. TBProfiler and Fastlin were found to agree on strain sublineage assignment in 99.4% of cases. As TBProfiler and Fastlin perform near identically in sublineage assignment and use the same underlying barcode, further comparisons are made between TBProfiler and TBtypeR only. TBtypeR agreed with TBProfiler in 57.2% of cases, gave more specific sublineage assignments for 23.5% of cases, and gave less specific sublineage assignments in 3.5% of cases. The remaining 15.8% of discordant cases were due to different strain nomenclature used in the respective SNP barcodes, largely due to: 1) the inclusion of additional lineage 2 strains in the TBtypeR SNP barcode from Thawornwattana et al^14^; and 2) TBtypeR using the new lineage names 1.3.1 and 1.3.2 instead of the older lineage names 1.2.2.1 and 1.2.2.2, as detailed in Napier et al^10^. TBtypeR and TBProfiler strain assignments agreed in 99.6% of cases when the TBtypeR SNP barcode was used with TBProfiler.

Finally, in order to demonstrate the extent to which mixed infections have been underreported due to the low sensitivity of existing tools to confidently detect low frequency mixtures, we ran TBtypeR (workflow FastTBtypeNF) on 5,000 samples randomly selected from the Wang et al study where mixed infections were screened for with TBProfiler^35^. Indeed, we found significantly higher levels of mixed infection, with a total of 6.1% (305/5000, 95% binomial confidence interval 5.5-6.8%) of samples being called mixed (Table S2), approximately 6-fold higher than the 1.1% reported by Wang et al. All mixture calls made by TBProfiler were also made by TBtypeR, with the majority (85.0%) of additional mixtures detected by TBtypeR being below 5% minor strain fraction (Figure 2A). This result suggests that the majority of mixed infections may constitute low-frequency mixtures which are difficult to detect with existing tools and highlights the importance of using more sensitive tools to capture the full genomic diversity of infecting strains. This is particularly relevant when using WGS for transmission mapping.

**Figure 2:**
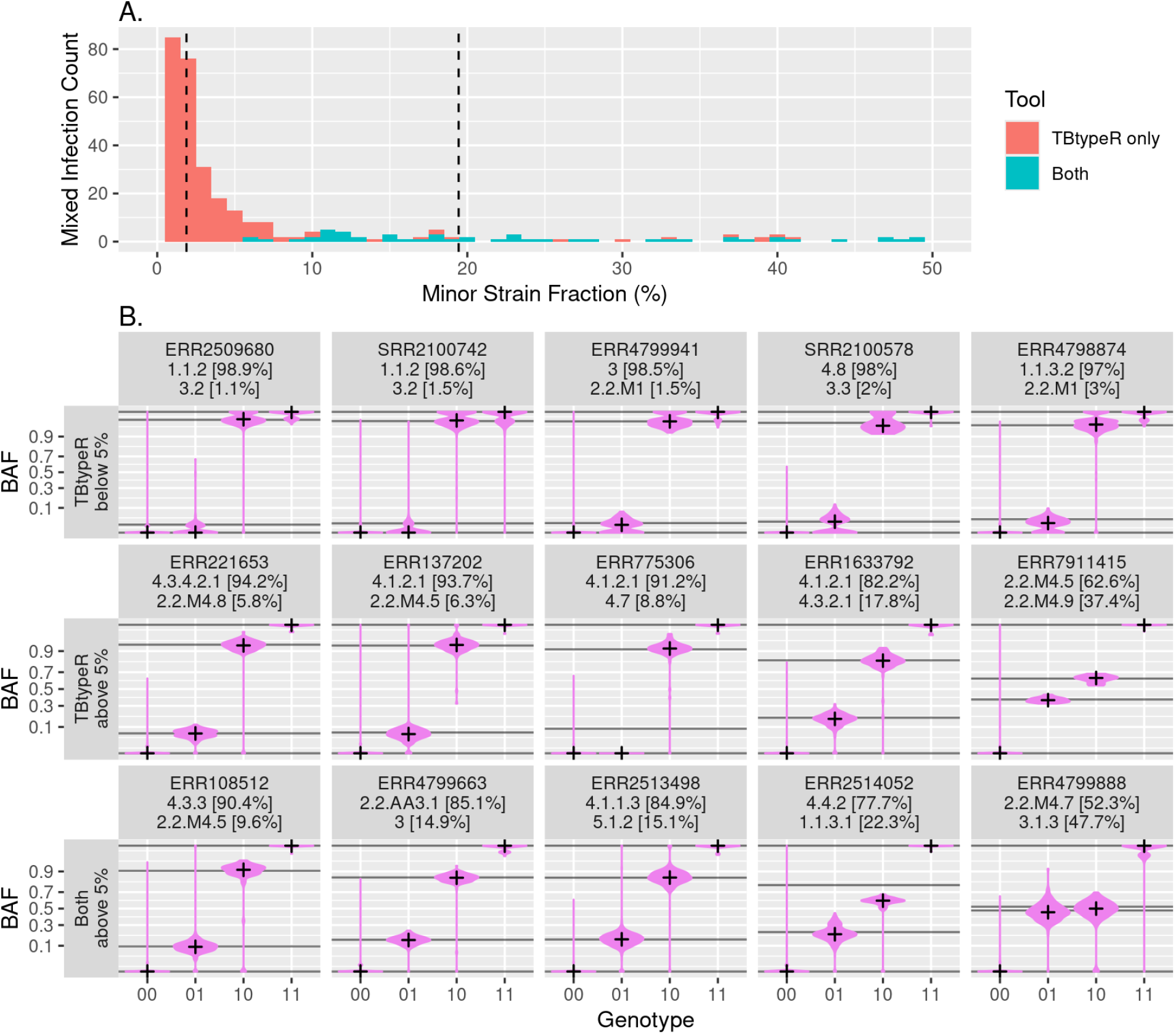
Comparison of TBtypeR and TBProfiler mixed infection calls from the Wang et al. dataset. A) Histogram of minor strain fraction of mixtures detected by TBtypeR and TBProfiler. Segments are coloured by whether the call was made by TBtypeR alone or both TBtypeR and TBProfiler. No calls were made by TBProfiler alone. Vertical dashed lines indicated the median minor strain fraction identified by TBtypeR (2.1%) and by TBProfiler (17.8%). The x-axis bins have a width of 1%. B) B-allele Frequency (BAF) violin plots showing BAF distribution vs TBtypeR predicted mixture genotypes. The top row shows 5 arbitrary mixtures detected by TBtypeR alone with below 5% minor strain fraction, the middle row shows 5 arbitrary mixtures detected by TBtypeR alone with above 5% minor strain fraction, and the bottom row shows 5 arbitrary mixtures detected by both TBtypeR and TBProfiler with above 5% minor strain fraction. Genotype indicates the major strain genotype followed by minor strain genotype, where ‘0’ indicates a reference allele and ‘1’ indicates an alternate allele. Horizontal lines indicate the expected B-allele frequency based on the minor strain fraction and crosses indicate the median observed B-allele frequency. The y-axis is arcsine transformed for better resolution of binomially distributed data. Labels indicate the WGS accession number as well as the sublineages and minor strains fractions identified. For simplicity TBtypeR was configured to call mixtures of at most 2 strains for this analysis, however ERR2514052 likely contains a third strain as evidence by the “01” genotype B-allele frequency being shifted lower than expected.

In order to demonstrate that the low-frequency mixtures detected by TBtypeR are likely to be genuine, we have visually compared the distribution of the B-allele frequency at the modelled mixture genotypes to the predicted minor strain fraction for a subset of samples (Figure 2). Five samples each were randomly selected covering cases where: 1) the predicted minor strain fraction is below 5% and the prediction is made by TBtypeR alone; 2) the predicted minor strain fraction is above 5% and the prediction is made by TBtypeR alone; and 3) the predicted minor strain fraction is above 5% and the prediction is made by both TBtypeR and TBProfiler. No mixtures with a minor strain fraction below 5% were predicted by TBProfiler. In all cases the B-allele frequency is shifted away from 0 (reference allele) and 1 (alternate allele) at sites with disagreeing mixture genotypes (“01” and “10”) across all barcode sites, with the magnitude of shift being consistent with the estimated minor strain fraction from TBtypeR.

## Discussion

Here we have presented TBtypeR, the most sensitive and accurate tool for identification of mixed infections from MTBC WGS data developed to date. TBtypeR has greater sensitivity and accuracy for detecting mixed infections at almost all tested parameters, with the widest performance margin at low frequency mixed infections where existing tools struggle. It also generally provides more specific sublineage classification than TBProfiler and can detect strains from 5 MTBC species covering 12 lineages. TBtypeR has been designed for and tested with short-read (Illumina) WGS data, but theoretically could be used with whole genome sequencing data from any platform (e.g. Oxford Nanopore long-read) if sufficient coverage is achieved. The provided SNP barcode is the most comprehensive to date, compiled and harmonized from five separate sources, and may be updated by users to improve sensitivity for emerging strains as new SNP markers become available.

Our benchmarking demonstrates that barcode-based tools outperform barcode-free tools for identification of MTBC mixed infections in most cases. We have shown that better performance is achieved by using SNP barcodes with as many sites as possible, with the performance of both TBProfiler and Fastlin improving when run with the 10-fold larger TBtypeR SNP barcode, and with only a minor increase in the respective runtimes. By utilising barcodes of known polymorphic sites, barcode-based tools are able to maximise the signal-to-noise ratio of the underlying data and thus yield more sensitive and accurate predictions of mixed infections. However, it is worth noting that barcode-free tools have the advantage that they may identify multiple strain infections in cases of either very closely related strains with small numbers of barcode SNPs differing, or with novel strains that are not yet captured by the SNP barcode. This limitation of barcode-based tools will be overcome by expanding the number of strains and SNPs included in the barcode as the knowledgebase of MTBC sublineages grows. We note however that the knowledgebase already has impressive geographic coverage with over 35,000 strains covering Africa, the Americas, Asia, Europe and Oceania used to derive the phylogenetic SNP barcode used (Table 1).

TBtypeR achieves improved performance over other tools for two key reasons: 1) use of an extensive SNP barcode (as discussed above); and 2), employing a rigorous statistical framework based on maximising the joint likelihood of the observed allele counts across all SNP sites under the binomial distribution. The binomial distribution is a much closer approximation of the underlying stochastic processes that drive the observed reference and alternate allele counts and naturally leads to more accurate estimates than the Gaussian mixture model employed by other tools. The difference between Gaussian and binomial models becomes particularly pronounced at both lower sequencing depths and lower minor strain fractions, when the probability of observing an alternate allele at any given polymorphic site becomes low.

We believe that our tool, TBtypeR, will be of use to the research community, and that new insights of epidemiological and clinical importance will be possible due to the ability to accurately identify low frequency mixed infections from WGS data. The full impact of mixed infections on diagnosis, treatment, and transmission mapping of TB will become evident with time. However, improved software solutions are only part of the equation when it comes to comprehensive detection of mixed infections. Moreno-Molina *et al* demonstrated that mixed infections are not uniformly distributed in resected lung tissue, and found a large discrepancy between the rates of mixture detection between sputum samples (5%) and lung tissue resection samples (39%)^36^. Part of this discrepancy may be because sputum samples are primarily composed of the proportion of bacteria in the body in closest proximity to the airway and in some cases may even derive from a biofilm lining a cavity. Consequently, the genetic diversity of bacteria in sputum can be limited. Secondly, all sample types are typically cultured prior to sequencing, thus biasing detection towards isolates that grow well under conventional culture conditions. As such, it has been demonstrated that greater genetic diversity is detected when performing direct sequencing of sputum samples compared to culture^37^. Taken together, we believe that careful selection of both software tools and sampling approaches will lead to increased routine detection of mixed infection from WGS data that more closely reflects the physiological truth found in individuals with TB.

## Data Availability

All data and source code produced are available online in the TBtypeR GitHub repository

https://github.com/bahlolab/TBtypeR

## Acknowledgements

This work was supported by a Walter and Eliza Hall Institute Innovation Fund grant to A.K.C and M.B. M.B. was supported by an NHMRC Investigator grant (GNT1195236). A.K.C was partially supported by an NHMRC Ideas grant (GNT2020750). Additional funding was provided by the NHMRC Independent Research Institute Infrastructure Support Scheme and the Victorian State Government Operational Infrastructure Program.

## Supplementary Figures

**Figure S1:**
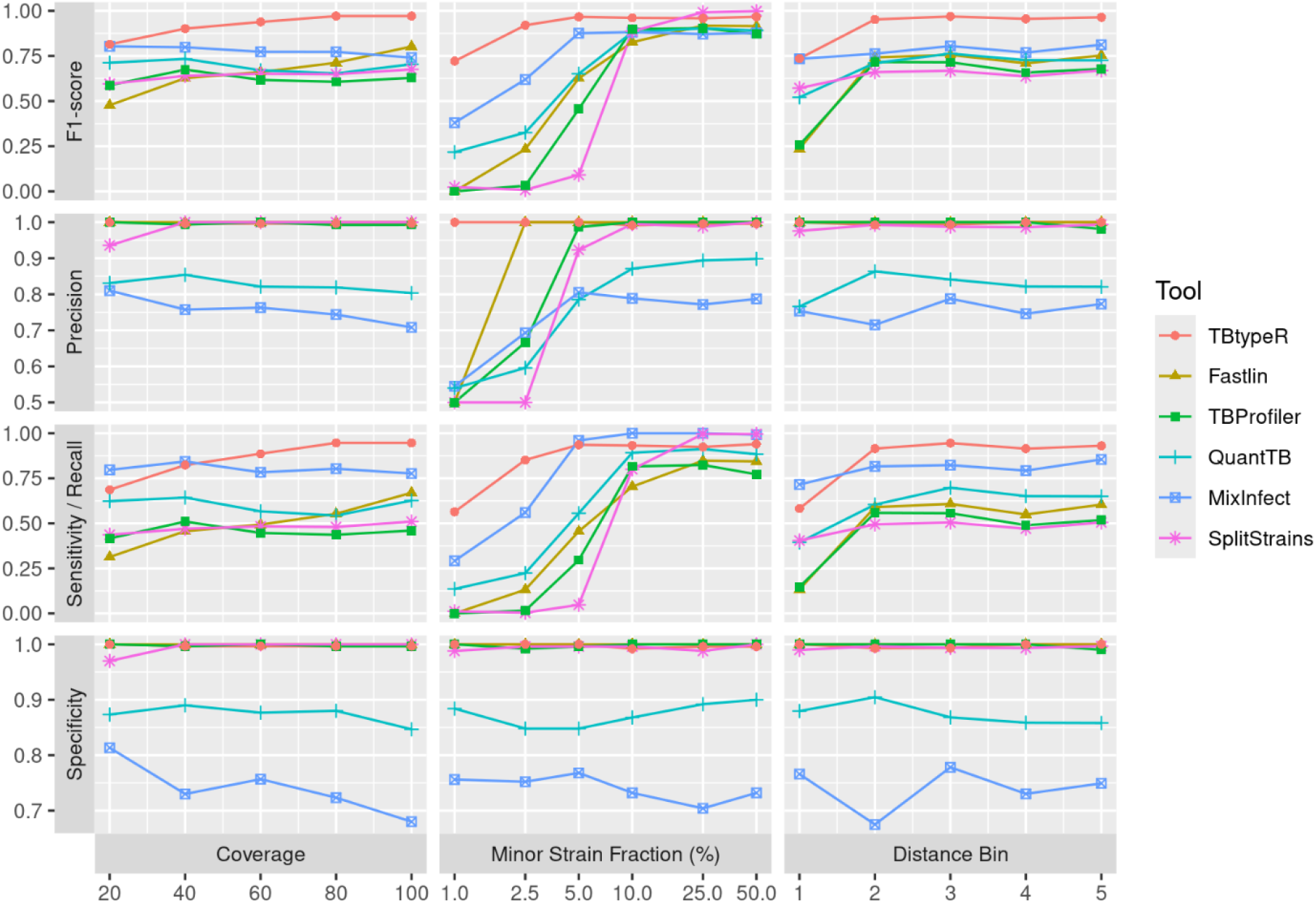
Additional binary performance metrics for each tool on the “in silico duos” dataset. The x-axis of minor strain fraction is log-2 scaled for improved resolution.

